# Strong immunogenicity of heterologous prime-boost immunizations with the experimental vaccine GRAd-COV2 and BNT162b2 or ChAdOx1-nCOV19

**DOI:** 10.1101/2021.06.22.21258961

**Authors:** Chiara Agrati, Stefania Capone, Concetta Castilletti, Eleonora Cimini, Giulia Matusali, Silvia Meschi, Eleonora Tartaglia, Roberto Camerini, Simone Lanini, Stefano Milleri, Stefano Colloca, Alessandra Vitelli, Antonella Folgori

## Abstract

Here we report on the humoral and cellular immune response in eight volunteers who autonomously chose to adhere to the Italian national COVID-19 vaccination campaign more than 3 months after receiving a single administration GRAd-COV2 vaccine candidate in the context of the phase 1 clinical trial. We observed a clear boost of both binding/neutralizing antibodies as well as T cell responses upon receipt of the heterologous BNT162b2 or ChAdOx1-nCOV19 vaccines. These results, despite the limitation of the small sample size, support the concept that a single-dose of an adenoviral vaccine may represent an ideal tool to effectively prime a balanced immune response, which can be boosted to high levels by a single dose of a different vaccine platform.

## Main Text

During the first year of the COVID-19 pandemic several clinical trials provided evidence of the efficacy of a number of candidate vaccine against SARS-CoV-2 ^1^. However, with new COVID-19 waves and global vaccine shortages the benefits of mixing two vaccines into one heterologous vaccination regimen are apparent and point towards a possibility of implementing this regimen to reach broad vaccine coverage ^2^. A heterologous vaccination regimen may also be needed to maintain protective immunity against SARS-CoV-2 over time. Finally, the use of a different vaccine has been suggested as a way to overcome hesitancy in persons who received a first dose of an adenoviral vectored vaccine for which safety concerns were raised. In fact, several European countries, such as Spain and Germany, are recommending a booster dose with mRNA vaccines to people under the age of 60 that received primary vaccination with Vaxzevria.

There is experimental evidence suggesting that heterologous vaccination improves the immune responses ^3^ and indeed this strategy has been proposed in vaccination against other viruses ^4^ and cancer ^5^. Preclinical data in mice confirm the immunological benefit of combining COVID-19 vaccines from different platforms ^6,7^ and several clinical trials are presently underway to test the safety and immunogenicity of this strategy for SARS-CoV2 infection.

We recently concluded a Phase 1 study to assess safety and immunogenicity of a single-dose regimen of a gorilla adenovirus vectored vaccine (GRAd-COV2) in younger (aged 18-55) and older (65-85) volunteers ^8^. The results showed good safety of the GRAd-COV2 vaccine and induction of both humoral (binding as well as neutralizing) and Th1 skewed cell-mediated immune response. At the final 24 weeks monitoring visit, eight enrolled volunteers reported that, outside the study procedures and in the context of the Italian National Vaccination Campaign (Table 1), they received either a full course (n=3) or only the first dose of BNT162b2-mRNA (n=3), or one dose of ChAdOx1 (n=2). These vaccinations occurred in a time frame between 14 and 24 weeks after the GRAd-COV2 study vaccination, and therefore side effects were not collected. These subjects provided blood samples to perform protocol defined laboratory test on anti-SARS-CoV2 humoral and cell-mediated immune response.

**Table 1.** Volunteers characteristics and vaccination history.

|  | **Volunteer id** | **gender/age range** | **study arm** | **GRAd-COV2 dose (vp\*)** | **Pfizer/AZ dose(s) @ study week** |
| --- | --- | --- | --- | --- | --- |
| **Pfizer x 2** | **A** | M/26-30y | 2B | 1 x 10<sup>11</sup> vp | week 16-19 |
|  | **B** | M/21-25y | 3B | 2 x 10<sup>11</sup> vp | week 19-22 |
|  | **C** | M/76-80y | 5B | 1 x 10<sup>11</sup> vp | week 14-17 |
| **Pfizer x 1** | **D** | M/71-75y | 4B | 5 x 10<sup>10</sup> vp | Pfizer x 1, week 22 |
|  | **E** | M/71-75y | 6B | 2 x 10<sup>11</sup> vp | Pfizer x 1, week 17† |
|  | **F** | M/21-25y | 2B | 1 x 10<sup>11</sup> vp | Pfizer x 1, week 24 |
| **ChAdOx1 x 1** | **G** | M/76-80y | 4B | 5 x 10<sup>10</sup> vp | AZ x 1, week 21 |
|  | **H** | M/71-75y | 5B | 1 x 10<sup>11</sup> vp | AZ x 1, week 21 |

\*vp= viral particles; †final study visit (w24) anticipated at week 20

In response to the heterologous vaccines, the levels of anti-Spike binding antibody levels measured at week 24 were greatly amplified compared to week 12, and exceeded the peak level recorded in each individual around week 4-8 post GRAd-COV2 vaccination (Figure 1), with no major differences in subjects receiving one or two doses of the heterologous vaccine. The only exception is volunteer F, who received the first BNT162b2 only 3 days before the week 24 visit, whose Spike Ab response slightly contracted from the last week 12 visit. Similarly, neutralizing titers at week 24 (MNA_90_) were potently boosted in all volunteers except F, and clearly exceeded those measured at study week 4. T cell responses were also assessed throughout the study visits by IFNγ ELISpot on frozen PBMC. Interestingly, the strong T cell response induced by GRAd-COV2 was further boosted with respect to both the peak and/or the week 12 levels in two volunteers receiving two mRNA doses (A and B), one receiving one mRNA dose (E) and in two volunteers receiving one ChAdOx1 dose (G and H). In the remaining subjects receiving one (D) or two mRNA doses (C) the T cell responses at week 24 were maintained at the same level as three months before, but did not further contract as observed in volunteer F.

**Figure 1.**
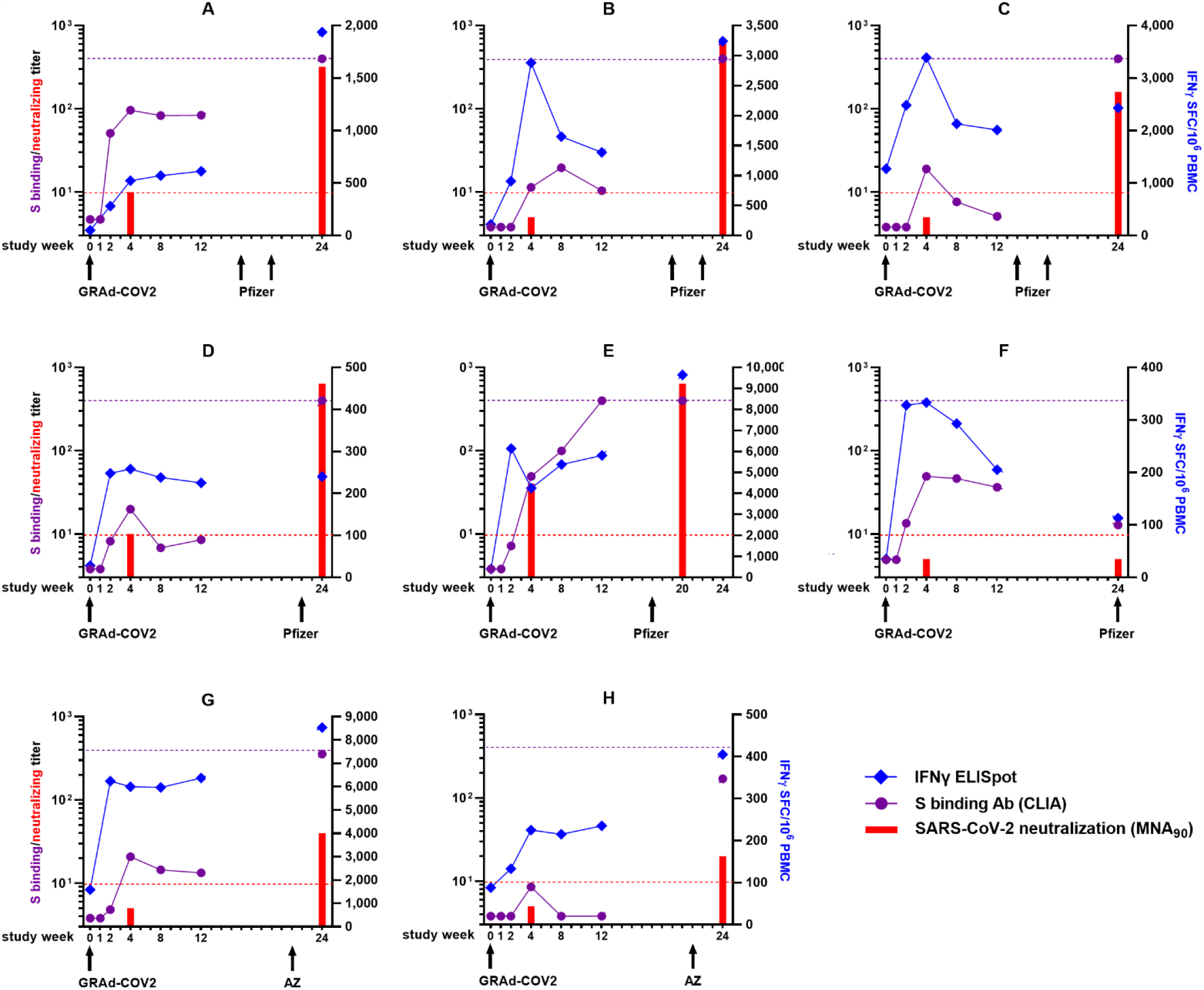
Immunogenicity profile of GRAd-COV2 vaccinated volunteers receiving approved COVID-19 vaccines. Each graph corresponds to an individual volunteer, as indicated by the code in the graph title. Arrows at the graph bottom indicate study week when each GRAd-COV2 or Pfizer (BNT162b2)/AZ (ChAdOx1-nCoV19) vaccination was received. Numbers on the x axis indicate timing in weeks from GRAd-COV2 vaccination when a study visit occurred. The purple round symbols indicate Spike binding IgG levels, as measured by CLIA assay and expressed in arbitrary units (AU)/ml. The red bars at w4 and 24 show SARS-CoV-2 neutralizing antibody titers, expressed as MNA_90_. Both serological endpoints are plotted against left y axis. Purple dotted line set at 400 AU/ml indicates upper limit of quantification for CLIA. Dotted red line indicates positivity threshold for the MNA assay, i.e. a neutralization titer of 1:10 or higher is deemed positive. The blue diamond symbols show Spike-specific T cell response as measured by IFNγ ELISpot, expressed as IFNγ Spot Forming Cells (SFC)/million PBMC, and plotted against right y axis.

This report has obvious limitations in the number of subjects and in the lack of safety data of the heterologous administration. Nevertheless, our observations suggest that the immune responses induced by the single-dose GRAd-COV2 vaccination can be effectively boosted also by a single heterologous vaccination with two approved vaccines, amplifying both antibody and T cell response.

These data are confirmatory of recently disclosed results from a study of safety and immunogenicity of homologous and heterologous immunization with ChAdOx1-nCoV19 and BNT162b2 ^9^.

Overall, these evidences support the concept that a single-dose of an adenoviral vaccine, which is cheap and easy to deploy in a pandemic setting, represents a good tool to effectively prime the immune system, which can be boosted afterwards by a single dose of a different vaccine platform, reaching high levels of immune responses. As preclinical and clinical evidence suggest ^6,9^, keeping adenoviral vectored vaccines as part of heterologous vaccination regimens may be the key for optimal induction and persistence of T cell responses, a feature that may confer cross-protection in the context of already circulating and newly emerging SARS-CoV-2 variants ^10^. Further prospective and controlled investigations are mandatory to confirm these data and to evaluate the impact of age and the optimal timing between prime and boost as well as the order of administration of the heterologous vaccines.

## Data Availability

All data generated or analyzed during this study are included in this published article

## Methods

### Study design

RT-CoV-2 is a phase 1, dose-escalation, open label clinical trial in two cohorts of healthy younger (18-55) or older (65-85) adults. Each age cohort consisted of 3 arms of 15 volunteers each, to assess safety and immunogenicity of a single GRAd-COV2 intramuscular administration at three different dose levels: 5×10^10^ viral particles (vp) (arms 1-younger and 4-older); 1×10^11^ vp (arms 2-younger and 5-older); 2×10^11^ vp (arms 3-younger and 6-older). All participants provided written informed consent before enrolment. The trial was conducted at the National Institute for Infectious Diseases Lazzaro Spallanzani (INMI) in Rome and at Centro Ricerche Cliniche in Verona (CRC-Verona), according to the Declaration of Helsinki, and approved by the Italian Regulatory Drug Agency (AIFA) and the Italian National Ethical Committee for COVID-19 clinical studies (ClinicalTrials.gov NCT04528641; EudraCT 2020-002835-31). Further information on study design and interim analysis safety and immunogenicity data can be found in ^8^. A phase 2 trial of GRAd-COV2 vaccine is currently ongoing (NCT04791423).

### SARS-CoV-2 anti-Spike IgG high throughput Chemiluminescence Immunoassay

Serum was collected at planned study visits the day of vaccination (week 0) and 1, 2, 4, 8, 12 and 24 weeks later. Spike binding IgG were measured using a chemiluminescence immunoassay (CLIA), namely DiaSorin LIAISON® SARS-CoV-2 S1/S2 IgG test on LIAISON® XL analyzers (DiaSorin, Italy), following manufacturer instructions. IgG concentrations are expressed as arbitrary units, AU/mL. According to datasheet, results >15 are clearly positive, between 12 and 15 are equivocal and <12 are negative or may indicate low level of IgG antibodies to the pathogen.

### SARS-CoV-2 microneutralization assay (MNA)

Neutralizing antibodies to SARS-CoV-2 were assessed by a microneutralization assay with live SARS-CoV-2 virus (strain 2019-nCoV/Italy-INMI1; GISAID accession ID: EPI_ISL_412974). The assay is described in detail in ^8^ and is based on inhibition of Vero E6 cells infection by serum dilution curves, with cytopathic effect (CPE) determination at 48h post infection. The highest serum dilution inhibiting at least 90% of the CPE was indicated as the neutralization titre and expressed as the reciprocal of serum dilution (MNA_90_). Briefly, heat-inactivated and titrated sera (duplicate two-fold serial dilutions, starting dilution 1:10) were mixed with equal volumes of 100 TCID_50_ SARS-CoV-2 and incubated at 37 °C 5% CO_2_ for 30 min. Subsequently, 96-well tissue culture plates with sub-confluent Vero E6 cell monolayers were infected with 100 μl/well of virus-serum mixtures and incubated at 37 °C and 5% CO2. To standardize inter-assay procedures, positive control samples showing high (1:160) and low (1:40) neutralizing activity were included in each MNA session. After 48 hours, microplates were observed by light microscope for the presence of CPE and then stained with Crystal Violet solution containing 2% Formaldehyde. Cell viability was measured by photometer at 595 nm (Synergy(tm) HTX Multi-Mode Microplate Reader, Biotek). The highest serum dilution inhibiting at least 90% of the CPE was indicated as the neutralization titre and expressed as the reciprocal of serum dilution (MNA_90_).

### IFNγ ELISpot assay on cryopreserved PBMC

The magnitude and kinetics of vaccine-induced Spike-specific T cells, as a function of gamma-interferon (IFNγ) production in response to antigen restimulation, was assessed by standard IFNγ ELISpot. Peripheral blood mononuclear cells (PBMC) were isolated by standard Histopaque (Sigma Aldrich) gradient technique the day of vaccination (week 0) and 2, 4, 8, 12 and 24 weeks later. Cryopreserved PBMC were thawed and rested overnight at 37°C in R10 medium [RPMI 1640 (Sigma Aldrich) supplemented with 10% heat-inactivated highly defined fetal bovine serum (FBS-HyClone), 2 mmol/L L-glutamine, 10 mmol/L HEPES buffer (N-2-hydroxyethylpiperazine-N-2-ethane sulfonic acid, Sigma Aldrich), 100 U/ml penicillin, and 100 µg/mL streptomycin (Gibco)]. Rested PBMC were plated at 2 × 10^5^ cells/well in ELISpot plates (Human IFN-γ ELISpot plus kit; Mabtech) and stimulated for 18-20 hours with 15mer peptides overlapping by 11 amino acids covering the full length Spike protein (synthetized by Elabscience Biotech Inc, distributed by TEMA RICERCA), arranged in 2 pools: S1 and S2 (3 μg/ml final concentration of each peptide). At the end of incubation, ELISpot assay was developed according to manufacturer’s instructions. Spontaneous cytokine production (background) was assessed by incubating PBMC with DMSO, the peptides diluent (Sigma). Results are expressed as spot forming cells (SFC)/10^6^ PBMCs in stimulating cultures, by summing responses to S1 and S2 after subtracting background.

## Data availability statement

All data generated or analyzed during this study are included in this published article

## Acknowledgments

We are grateful to Dr. Enrico Girardi (INMI Lazzaro Spallanzani IRCCS) for insightful discussion and critical reading of the manuscript.

The phase 1 study has been funded by Regione Lazio and Italian Ministry of research.

INMI authors are supported by the Italian Ministry of Health (Ricerca Corrente line 1, COVID-2020-12371735 and COVID-2020- 12371817).

## Author contributions

Conceptualization: C.A., S.C., C.C., S.L., R.C., S.Co., A.F., Investigation: C.A., C.C., E.C., G.M., S.Me., E.T., S.M., S.L. Writing – original draft: C.A., S.C., A.V. Writing – review & editing: all authors.

